# Comparison of COVID-19 Infections Among Healthcare Workers and Non-Healthcare Workers

**DOI:** 10.1101/2020.08.13.20174482

**Authors:** Rachel Kim, Sharon Nachman, Rafael Fernandes, Kristen Meyers, Maria Taylor, Debra LeBlanc, Adam J Singer

## Abstract

**Objectives:** Healthcare workers face distinct occupational challenges that affect their personal health, especially during a pandemic. In this study we compare the characteristics and outcomes of Covid-19 patients who are and who are not healthcare workers (HCW).

**Methods:** We retrospectively analyzed a cohort of adult patients with known HCW status and a positive SARS-CoV-2 PCR test presenting to a large academic medical center emergency department (ED) in New York State. We routinely collect data on occupation and exposures to suspected Covid-19. The primary outcome was hospital admission. Secondary outcomes were ICU admission, need for invasive mechanical ventilation (IMV), and mortality. We compared baseline characteristics and outcomes of Covid-19 adult patients based on whether they were or were not HCW using univariable and multivariable analyses.

**Results:** From March 1 2020 through June 2020, 2,842 adult patients (mean age 53+/-19 years, 53% male) with positive SARS-CoV-2 PCR tests and known HCW status visited the ED. This included 193 (6.8%) known HCWs and 2,649 (93.2%) non-HCWs. Compared with non-HCW, HCWs were younger (43 vs 53 years, P<0.001), more likely female (118/193 vs 1211/2649, P<0.001), and more likely to have a known Covid-19 exposure (161/193 vs 946/2649, P<0.001), but had fewer comorbidities. On presentation to the ED, HCW also had lower frequencies of tachypnea (12/193 vs 426/2649, P<0.01), hypoxemia (15/193 vs 564/2649, P<0.01), bilateral opacities on imaging (38/193 vs 1189/2649, P<0.001), and lymphocytopenia (6/193 vs 532/2649, P<0.01) compared to non-HCWs. Direct discharges home from the ED were more frequent in HCW 154/193: 80% vs 1275/2649: 48% p<0.001). Hospital admissions (38/193 20% vs 1264/2694 47%, P<0.001), ICU admissions (7/193 3% vs 321/2694 12%, P<0.001), need for IMV (6/193 3% vs 321/2694 12%, P<0.001) and mortality (2/193 1% vs 219/2694 8%, P<0.01) were lower than among non-HCW. After controlling for age, sex, comorbidities, presenting vital signs and radiographic imaging, HCW were less likely to be admitted (OR 0.6, 95%CI 0.3-0.9) than non HCW.

**Conclusions:** Compared with non HCW, HCW with Covid-19 were younger, had less severe illness, and were less likely to be admitted.

## Introduction

Illness in healthcare workers (HCWs) is of particular concern in light of the coronavirus disease 19 (Covid-19) pandemic. HCWs are often at risk for increased exposure to infectious diseases compared to the general population, and they may serve as vectors for transmission of these illnesses [1]. The repeated exposure to pathogens may also alter the course of infectious disease in HCWs compared to that in the general population. There is currently little literature studying how HCWs with Covid-19 have fared, and whether their course of disease may be different than non-HCWs.

In April 2020 a CDC Morbidity and Mortality Report documented that HCWs made up 3% of COVID-19 cases in the US, however HCW status was only reported in 16% of cases [2]. In states that reported HCW status of all Covid-19 cases, HCWs made up 11% of cases [2]. In our hospital, routine documentation of healthcare worker status and exposure to patients with suspected Covid-19 was added to the emergency department (ED) patient intake form at the end of February.

The objectives of the current study were to compare baseline characteristics and subsequent outcomes in adult HCW or non HCW patients with Covid-19 presenting to a large ED, including those admitted to our hospital and those directly discharged from the ED.

## Methods

### Study Design

We conducted a structured, retrospective review of all patients presenting to our ED with confirmed Covid-19 based on a positive SARS-CoV-2 PCR. This study followed the Strengthening of Reporting of Observational Studies in Epidemiology (STROBE) reporting guidelines for cross-sectional studies [3]. We also followed the recommended methodology of Kaji et al. for retrospective chart reviews [4]. We received IRB approval with waiver of informed consent due to the retrospective nature of this study.

### Patients and Setting

Our hospital is a 650 bed, suburban academic tertiary-care hospital located approximately 60 miles from Manhattan. Patient volume is approximately 110,000 adult and pediatric patients per year. Eligible patients presented from March 1, 2020 to June 30, 2020 and included all patients over 18 years of age with Covid-19 confirmed by a positive SARS-Cov-2 PCR. In our ED we routinely collected data on patient occupation and exposure to suspected Covid-19 throughout the study period. Patients without information regarding whether they were HCW or not were excluded.

### Data Source and Collection

Data included in our analysis were either automatically extracted from the electronic medical record (e.g., vital signs and laboratory results) or were abstracted through manual chart review (e.g., symptoms and HCW status). Specific data extracted included demographics, past medical history and comorbidities based on patient report, baseline vital signs, symptoms at presentation, laboratory test results, radiological findings, treatments, and outcomes (e.g., disposition, need for IMV, and mortality).

We defined all study data and variables prior to initiating the study and trained our data abstractors using a library of definitions. Healthcare workers included physicians, nurses, technicians and any other support staff that come into direct contact with patients. Hypoxemia was defined as an oxygen saturation less than 94% while tachypnea was defined as a respiratory rate greater than 24/min. We periodically monitored data collection and determined the inter-observer agreement on the primary and secondary outcomes on a randomly selected sample of 20 study patients. Interobserver agreement (Kappa statistic) ranged from 0.89 (95% CI, 0.67-0.99) to 1.0.

### Study Outcomes

The primary outcome was hospital admission. Secondary outcomes were ICU admission, need for invasive mechanical ventilation, and mortality.

### Data Analysis

Discrete data are summarized as numbers and frequencies for nominal data and compared with Chi-square or Fisher’s exact tests (when sample sizes were less than 5). Continuous data are summarized as means with standard deviations (SD) or medians with interquartile (IQR) ranges and compared with T-tests or Mann Whitney U tests for normally distributed and skewed data respectively. For all variables and models, we only used the initial findings at ED presentation. Exploratory multivariable analysis of primary and secondary outcomes was performed using potential predictor variables chosen based on biological plausibility and previous reports such as age, sex, comorbidities, presenting vital signs, chest XR findings, and HCW status. Comparisons between groups were performed using logistic and linear regression for binary and continuous data respectively. Level of significance was defined as a P value of 0.05 or less. Data was analyzed using SPSS for Windows version 27 (IBM, Armonk, NY).

## Results

### General Characteristics

From March 1 to June 30 2020 there were 2,888 Covid-19 infected patients that presented to our ED, of which 2,846 were over 18 years of age. HCW status was unknown in four patients leaving a sample size of 2,842 study patients. Overall, the mean (SD) age for adult patients was 53 (19) years, 1,513 (53%) were male and 1,306 (46%) were white. The total number of patients admitted to the hospital was 1,406 (49%) of which 328 (12%) were admitted to an ICU (either immediately or subsequent to admission to a regular floor). Invasive mechanical ventilation was required in 261/2842 (9.1%) patients and 221/2842 patients (7.8%) died.

### Univariable comparisons

A comparison of patient baseline characteristics based on HCW status is presented in Table 1. Compared with non HCW, HCW were younger (43+/-13 vs 53+/-18 years; mean difference 10, 95%CI 8 to 23, P<0.001] more likely female (61% vs. 46%; mean difference 15%, 95%CI 8-23, P<0.001), and less likely Hispanic (12% vs. 36%; mean difference 23%, 95%CI 17-28, P<0.001). HCW were less likely to have comorbidities (33% vs 49%; mean difference 16%, 95%%CI 8-22, P<0.01) but more likely to have reported a known contact with a person with Covid-19 symptoms (83% vs 36%; mean difference 48%, 95%CI 41-53, P<0.001) than non-HCW. While no HCW resided in a skilled nursing facility, 11% of non HCW came from a nursing home.

**Table 1.**
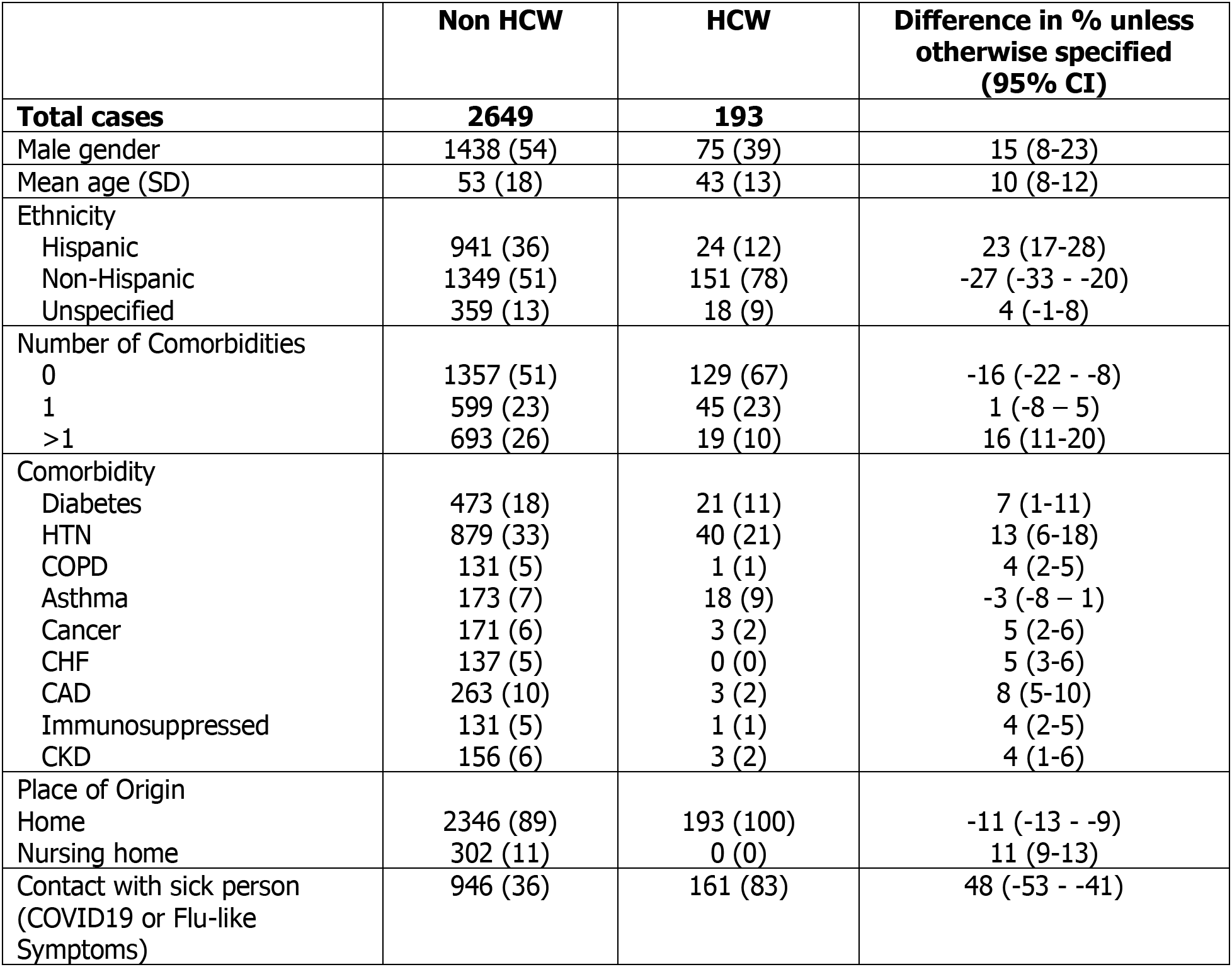
Patients General Characteristics.

|  | Non HCW | HCW | Difference in % unless otherwise specified (95% CI) |
| --- | --- | --- | --- |
| **Total cases** | **2649** | **193** |  |
| Male gender | 1438 (54) | 75 (39) | 15 (8-23) |
| Mean age (SD) | 53 (18) | 43 (13) | 10 (8-12) |
| Ethnicity |  |  |  |
| Hispanic | 941 (36) | 24 (12) | 23 (17-28) |
| Non-Hispanic | 1349 (51) | 151 (78) | -27 (-33 - -20) |
| Unspecified | 359 (13) | 18 (9) | 4 (-1-8) |
| Number of Comorbidities |  |  |  |
| 0 | 1357 (51) | 129 (67) | -16 (-22 - -8) |
| 1 | 599 (23) | 45 (23) | 1 (-8 - 5) |
| >1 | 693 (26) | 19 (10) | 16 (11-20) |
| Comorbidity |  |  |  |
| Diabetes | 473 (18) | 21 (11) | 7 (1-11) |
| HTN | 879 (33) | 40 (21) | 13 (6-18) |
| COPD | 131 (5) | 1 (1) | 4 (2-5) |
| Asthma | 173 (7) | 18 (9) | -3 (-8 - 1) |
| Cancer | 171 (6) | 3 (2) | 5 (2-6) |
| CHF | 137 (5) | 0 (0) | 5 (3-6) |
| CAD | 263 (10) | 3 (2) | 8 (5-10) |
| Immunosuppressed | 131 (5) | 1 (1) | 4 (2-5) |
| CKD | 156 (6) | 3 (2) | 4 (1-6) |
| Place of Origin |  |  |  |
| Home | 2346 (89) | 193 (100) | -11 (-13 - -9) |
| Nursing home | 302 (11) | 0 (0) | 11 (9-13) |
| Contact with sick person (COVID19 or Flu-like Symptoms) | 946 (36) | 161 (83) | 48 (-53 - -41) |

A comparison of initial vital signs, laboratory results and radiological findings based on HCW status is presented in Table 2. Compared with non HCW, HCW were less likely to be tachypneic (6% vs 13%; mean difference 7, 95%CI 2-10, P<0.01), hypoxemic (8% vs 22%; mean difference 13%, 95%CI 8-17, P<0.01), have bilateral lung opacities on imaging (20% vs 45%; mean difference 25, 95%CI 18-31, P<0.01), or have lymphopenia (6% vs 20%; mean difference 14, 95%CI 9-17, P<0.01).

**Table 2.**
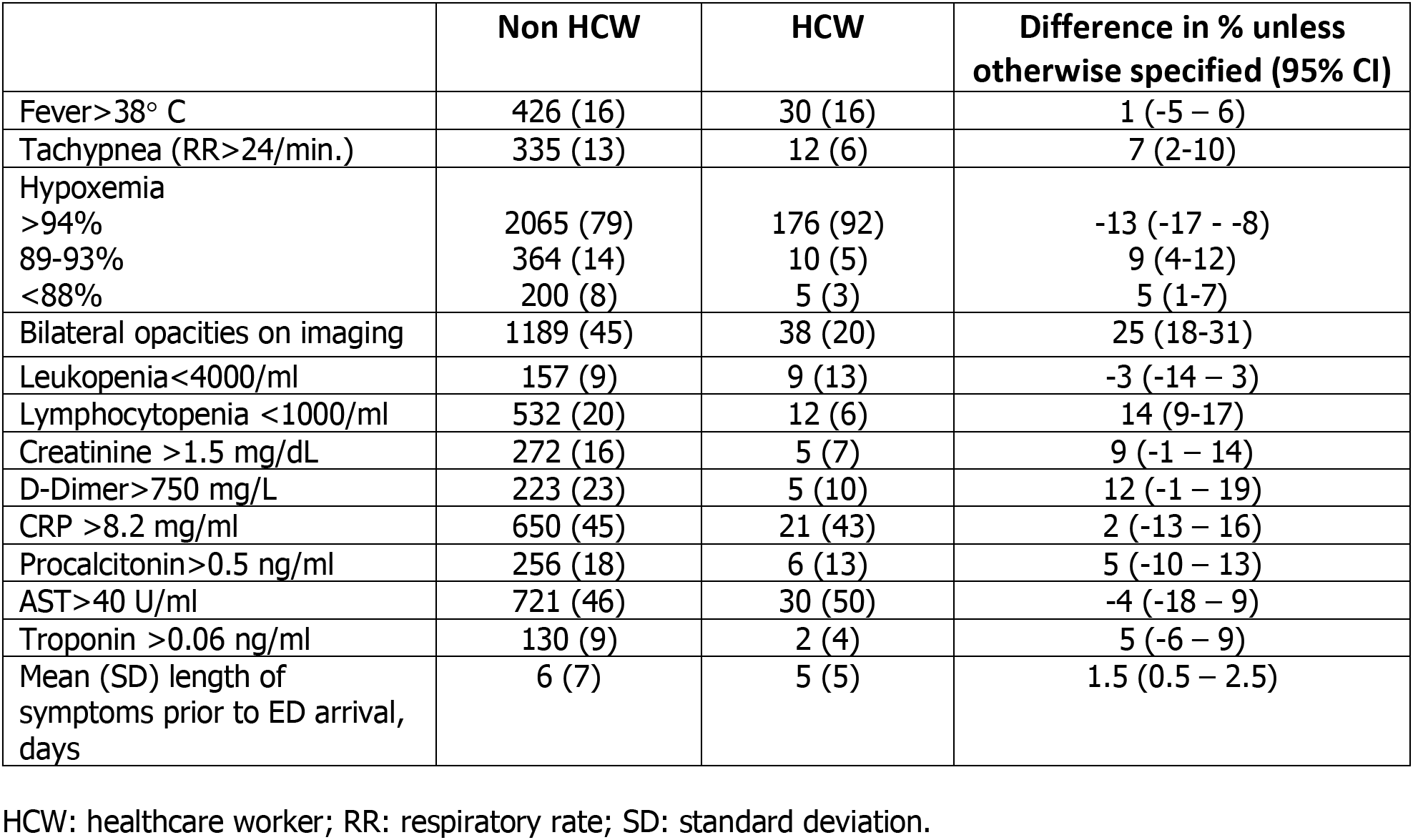
Signs, Imaging and Laboratory Characteristics on Presentation.

|  | Non HCW | HCW | Difference in % unless otherwise specified (95% CI) |
| --- | --- | --- | --- |
| Fever>38° C | 426 (16) | 30 (16) | 1 (-5 – 6) |
| Tachypnea (RR>24/min.) | 335 (13) | 12 (6) | 7 (2-10) |
| Hypoxemia |  |  |  |
| >94% | 2065 (79) | 176 (92) | -13 (-17 - -8) |
| 89-93% | 364 (14) | 10 (5) | 9 (4-12) |
| <88% | 200 (8) | 5 (3) | 5 (1-7) |
| Bilateral opacities on imaging | 1189 (45) | 38 (20) | 25 (18-31) |
| Leukopenia<4000/ml | 157 (9) | 9 (13) | -3 (-14 – 3) |
| Lymphocytopenia <1000/ml | 532 (20) | 12 (6) | 14 (9-17) |
| Creatinine >1.5 mg/dL | 272 (16) | 5 (7) | 9 (-1 – 14) |
| D-Dimer>750 mg/L | 223 (23) | 5 (10) | 12 (-1 – 19) |
| CRP >8.2 mg/ml | 650 (45) | 21 (43) | 2 (-13 – 16) |
| Procalcitonin>0.5 ng/ml | 256 (18) | 6 (13) | 5 (-10 – 13) |
| AST>40 U/ml | 721 (46) | 30 (50) | -4 (-18 – 9) |
| Troponin >0.06 ng/ml | 130 (9) | 2 (4) | 5 (-6 – 9) |
| Mean (SD) length of symptoms prior to ED arrival, days | 6 (7) | 5 (5) | 1.5 (0.5 – 2.5) |

HCW: healthcare worker; RR: respiratory rate; SD: standard deviation.

HCW were more likely than non HCW to be directly discharged from the ED (154 /193 [80%] vs 1275/2649 [48%]; mean difference 32, 95%CI 25-37 [P<0.001]). By comparison, non HCW were more likely to require ICU admission (321/2649 [12%] vs 7/193 [4%]; mean difference 8, 95%CI 4-11, P<0.001), invasive mechanical ventilation (6/193 [10%] vs 255/2649 [3%]; mean difference 7, 95%CI 3-9, P<0.01), and to die (2/193 [8%] vs 219/2649 [1%]; mean difference 7, 95%CI 4-9, P<0.001) than HCW. See table 3 for complete results.

**Table 3.**
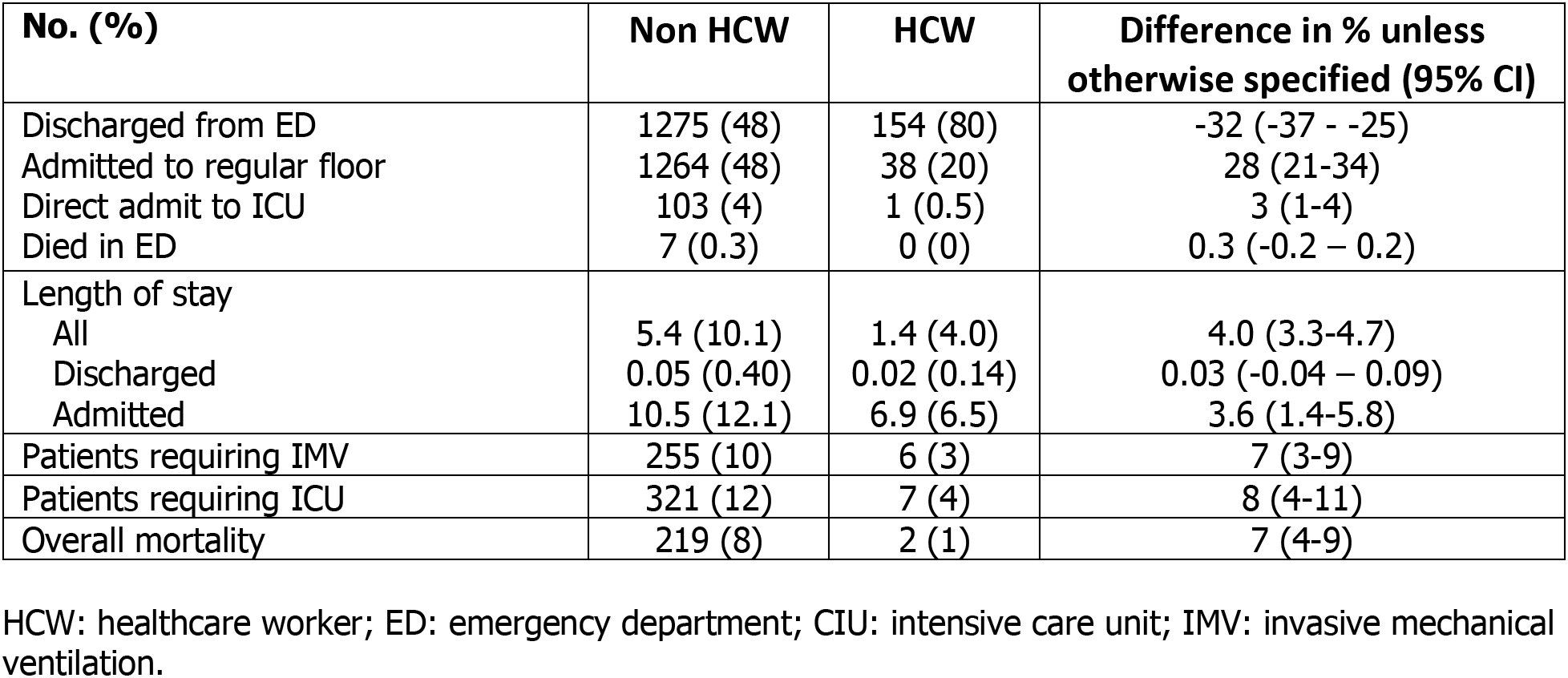
Disposition.

### Multivariable Analyses

After controlling for known potential confounders (including age, sex, comorbidities, vital signs, and CXR findings), variables significantly associated with admission included age (OR 1.05/year, 95%CI 1.04-1.06), fever (OR 2.1, 95%CI 1.57-2.81), tachypnea (OR 6.0, 95%CI 3.56-1.53), hypoxemia (OR 1.46, 95%CI 1.39-1.53) and presence of comorbidities (OR 1.75, 95%CI 1.35-2.25 and 4.1, 95%CI 3.0-5.5 for one or more comorbidities respectively). HCW were less likely than non HCW to require hospital admission (OR 0.58, 95%CI 0.36-0.92) even after adjusting for confounders (Table 4).

**Table 4.** Multivariable Predictors of Outcomes.

|  | OR | 95% CI |
| --- | --- | --- |
| **Hospital Admission** |  |  |
| Male sex | 1.09 | 0.88-1.35 |
| Age per year | 1.05 | 1.04-1.06 |
| Non-HCW | Reference | - |
| HCW | 0.58 | 0.36-0.92 |
| Temp > 38° C | 2.10 | 1.57-2.81 |
| Tachypnea >24/min. | 6.00 | 3.56-10.11 |
| Hypoxemia <94% | 1.46 | 1.39-1.53 |
| Any exposure | 0.50 | 0.40-0.63 |
| # comorbidities |  |  |
| 0 | Reference | - |
| 1 | 1.75 | 1.35-2.25 |
| 2+ | 4.06 | 3.01-5.47 |
| **Invasive Mechanical Ventilation** |  |  |
| Male | 2.04 | 1.48-2.81 |
| Age per year | 0.99 | 0.98-1.00 |
| Non-HCW | Reference | - |
| HCW | 0.97 | 0.38-2.45 |
| Temp > 38° C | 1.30 | 0.92-1.83 |
| Tachypnea > 24/min. | 1.24 | 0.96-1.78 |
| Hypoxemia <94% | 1.09 | 1.06-1.12 |
| Any exposure | 1.22 | 0.88-1.68 |
| # comorbidities |  |  |
| 0 | Reference | - |
| 1 | 1.36 | 0.91-2.04 |
| 2+ | 1.30 | 0.87-1/93 |
| **ICU Admission** |  |  |
| Male sex | 1.73 | 1.30-2.29 |
| Age per year | 0.99 | 0.98-1.00 |
| Non-HCW | Reference | - |
| HCW | 0.80 | 0.33-1.91 |
| Temp > 38° C | 1.16 | 0.85-1.60 |
| Tachypnea >24/min. | 1.54 | 1.11-2.13 |
| Hypoxemia | 1.08 | 1.05-1.10 |
| Any exposure | 1.13 | 0.84-1.52 |
| # comorbidities |  |  |
| 0 | Reference | - |
| 1 | 1.32 | 0.91-1.90 |
| 2+ | 1.40 | 0.97-2.00 |
| **Mortality** |  |  |
| Male | 1.97 | 1.38-2.82 |
| Age per year | 1.05 | 1.04-1.07 |
| Non-HCW | Reference | - |
| HCW | 0.94 | 0.21-4.24 |
| Temp > 38° C | 0.77 | 0.48-1.22 |
| Tachypnea >24/min. | 1.45 | 0.97-2.16 |
| Hypoxemia <94% | 1.09 | 1.06-1.12 |
| Any exposure | 0.80 | 0.54-1.19 |
| # comorbidities |  |  |
| 0 | Reference | - |
| 1 | 1.03 | 0.61-1.76 |
| 2+ | 1.50 | 0.94-2.41 |

Variables associated with ICU admission after adjusting for confounders were male sex (OR 1.73, 95%CI, 1.3-2.3), tachypnea (OR 1.54, 95%CI 1.1-2.13), and hypoxemia (OR 1.08, 95%CI 1.05-1.10). Variables associated with IMV were male sex (OR 2.04, 95%CI, 1.48-2.81) and hypoxemia (OR 1.09, 95%CI, 1.06-1.12). Variables independently associated with mortality included male sex (OR 1.97, 95%CI 1.38-2.82), age (OR 1.05/year, 95%CI 1.04-1.07) and hypoxemia (OR 1.09, 95%CI 1.06-1.12). Being a HCW was not associated with increased risk of ICU admission, IMV, or mortality.

## Discussion

Our study identifies clear differences in HCWs with Covid-19 presenting to the emergency department compared to non-HCWs. HCW were generally younger and with fewer comorbidities than non HCW. Compared with non HCW, HCW were less severely ill as evidenced by lower rates of tachypnea, hypoxemia, lymphopenia and the presence of bilateral opacities on radiographic imaging. As a result, HCW were less likely to be admitted to the hospital, to require ICU level of care, require invasive mechanical ventilation and to ultimately die than non HCW. This seemingly milder course of disease in HCWs compared to the general population is reported in other studies of Covid-19 in HCWs [5]. Given the large number of HCW in our catchment area and their potential high risk of exposure to Covid-19 [6], it is reassuring to find low rates of severe illness when compared to the general population. In our study we found that being a HCW was not associated with poor outcomes even after adjusting for age and comorbidities.

Certain characteristics of HCWs with COVID-19 may be attributed to common characteristics of HCWs in general. Although men constitute a larger percentage of total HCWs in the United States, women make up the majority of most types of healthcare occupations including registered nurses, home health aides, medical assistants, and physician assistants [7]. This may be reflected by the large majority of HCW Covid-19 cases in female HCWs. The higher frequency of HCW Covid-19 infection in females and those in the age group 16-44 has been repeatedly found in most studies that report this demographic information [2,5,8-15].

Overall, morbidity and mortality studies for HCWs are sparse, however those that exist suggest that HCWs, particularly physicians, tend to live longer than age and sex matched non HCW [16]. While some studies have found that physicians are more likely to have chronic conditions like obesity, metabolic syndromes, hypertension and cardiovascular disease, others have found that they are at lower risk for these illnesses [1]. HCWs are also more likely to have come in contact with a sick person because of the nature of their job [16]. This strongly supports the importance of continued use of appropriate PPE for all HCW. All of these factors have the potential to contribute to an altered course of Covid-19 in HCWs.

Our study has several notable limitations. Most importantly, our study was retrospective and subject to considerable selection bias and residual confounding. In addition, the number of healthcare workers was relatively limited. We also did not distinguish between various types of healthcare workers based on their specific profession. Furthermore, our study is representative of a single large academic center near the epicenter of the Covid-19 pandemic and may be less representative of other centers. We also did not control for ethnicity.

## Conclusions

In summary, we found that when compared to adult non-healthcare workers with Covid-19 presenting to the ED, healthcare workers with Covid-19 were more likely to have an identified COVID exposure, present less severely ill, and less likely to be admitted to the hospital. Longer term follow-up of HCW will be needed to better assess the risk of HCW.

## Data Availability

Will be made available after de-identifcation

## Acknowledgments

Research reported in this publication was supported by the Stony Brook University’s Renaissance School of Medicine COVID-19 Data Repository Quality Initiative instituted by the Office of the Dean of the Renaissance School of Medicine and supported by the Department of Biomedical Informatics.

